# Diffusion and Perfusion Heterogeneity for Survival Stratification in Post-Treatment Glioblastoma

**DOI:** 10.64898/2026.08.01.26359454

**Authors:** Yiğit Hasan Arı

## Abstract

**Purpose:** The prognostic value of diffusion- and perfusion-derived tumor-mask heterogeneity for overall survival in post-treatment glioblastoma was evaluated using a public MRI dataset.

**Materials and Methods:** The University of California San Diego Post-Treatment Glioblastoma (UCSD-PTGBM) dataset was used to construct a first-timepoint cohort of 133 subjects. Twenty tumor-mask features were extracted from high b-value apparent diffusion coefficient (ADC) and dynamic susceptibility contrast (DSC) perfusion maps. Prognostic associations were assessed using univariate and adjusted Cox regression. A benchmark compared clinical, diffusion, perfusion, and combined models using cross-validated concordance indices and permutation testing.

**Results:** ADC standard deviation (*ADC*_std_) showed the strongest univariate prognostic association (hazard ratio 1.56, false discovery rate *q* = 0.0003, concordance index 0.621) and remained independently significant after clinical adjustment (HR 1.48, *p* < 0.001). Mean transit time standard deviation (*MTT*_std_) was the strongest perfusion-derived feature (HR 1.38, *q* = 0.025, concordance index 0.578). *ADC*_std_ and *MTT*_std_ showed low correlation (Spearman *r* = 0.24). In cross-validation, neither imaging feature alone significantly improved discrimination over the clinical baseline (clinical plus ADC, ΔC = +0.058, *p* = 0.071; clinical plus MTT, ΔC = +0.035, *p* = 0.194). Only the model combining clinical variables, *ADC*_std_ and *MTT*_std_ achieved a significant improvement (concordance index 0.619; ΔC = +0.072, *p* = 0.029).

**Conclusion:** ADC heterogeneity was the numerically strongest imaging signal, while DSC perfusion heterogeneity was weaker and less consistent. Only the combined model significantly outperformed the clinical baseline, but not ADC alone, leaving perfusion’s contribution unproven.

## 1. Introduction

Glioblastoma (GBM) is the most common primary malignant brain tumor in adults and remains associated with poor survival despite multimodal treatment with maximal safe resection, concurrent chemoradiotherapy, and adjuvant temozolomide [1]. Although molecular factors including MGMT promoter methylation influence outcome [2], imaging-based survival stratification has been increasingly investigated through radiomics [3], volumetric assessment [4], and related prognostic modeling approaches. In the post-treatment setting, however, accurate prognostic assessment is particularly challenging because conventional MRI is frequently confounded by treatment-related changes, including pseudoprogression, radiation necrosis, and anti-angiogenic treatment effects [5]. These processes may mimic or obscure viable tumor and complicate image interpretation under established and updated Response Assessment in Neuro-Oncology (RANO) frameworks [6,7].

Diffusion MRI and dynamic susceptibility contrast (DSC) perfusion MRI provide complementary but biologically distinct information in treated GBM. Diffusion-derived measures reflect tissue microstructure and may capture intratumoral heterogeneity related to residual hypercellular tumor, necrosis, edema, and treatment-induced architectural disruption. In particular, heterogeneity on apparent diffusion coefficient (ADC) maps has emerged as a pragmatic imaging signal because ADC is widely available in routine neuro-oncologic MRI protocols. Histogram-based ADC measures have shown prognostic associations in both newly diagnosed and recurrent glioblastoma cohorts, particularly in bevacizumab-treated settings [8,9].

By contrast, DSC-derived metrics such as cerebral blood volume, cerebral blood flow, and mean transit time are widely utilized as surrogate markers of tumor vascularity and hemodynamic activity [10]. However, perfusion measurements are inherently complex in the post-treatment environment. They can be heavily confounded by blood-brain barrier disruption and by anti-angiogenic treatment-induced vascular changes, the latter of which have themselves been proposed as early markers of treatment response [5,11], as well as by variations in leakage correction and post-processing methodologies [10,12]. Consequently, although both modalities are biologically relevant, their relative prognostic utility within a simple, clinically accessible tumor-mask framework remains unclear.

The University of California San Diego Post-Treatment Glioblastoma (UCSD-PTGBM) dataset provides a public post-treatment cohort with multimodal MRI, neuroradiologist-approved tumor segmentations, and associated survival information [13]. Unlike most widely used public glioma resources, which primarily emphasize the preoperative setting, this dataset was specifically assembled for post-treatment imaging research and includes both diffusion and DSC perfusion imaging. In the present study, a focused comparison of diffusion- and perfusion-derived tumor-mask heterogeneity metrics was performed for scan-anchored overall survival prediction, utilizing the earliest eligible post-treatment MRI per subject. Incremental value beyond a simple clinical baseline was assessed using Cox modeling and cross-validated concordance analysis, with emphasis placed on whether conventional ADC heterogeneity or DSC perfusion heterogeneity provided the more informative prognostic signal in this cohort.

## 2. Methods

### 2.1. Dataset and Patient Population

The publicly available University of California San Diego Post-Treatment Glioblastoma (UCSD-PTGBM) dataset was accessed through The Cancer Imaging Archive (TCIA) [13,14]. The original descriptor reports 243 imaging timepoints from 178 subjects with histopathologically confirmed glioblastoma [13]. At the time of analysis, the publicly accessible imaging partition contained 184 studies from 136 subjects, together with the version 3 clinical spreadsheet in which previously misassigned patient identifiers had been corrected [14]. MRI examinations were acquired on 3 Tesla GE scanners at two UCSD-affiliated institutions and included standard anatomical imaging, diffusion imaging, DSC perfusion imaging, and neuroradiologist-approved voxelwise tumor segmentations [13]. The dataset is distributed through TCIA under a CC BY 4.0 license [14]. Original data collection was approved by the University of California San Diego Institutional Review Board (IRB #809620); no additional ethics approval was required for this secondary analysis of a de-identified public dataset [13].

### 2.2. Cohort Definition and Survival Endpoint

The original dataset publication described an overall survival subset restricted to scans acquired within 90 days after surgery that additionally included Restriction Spectrum Imaging (RSI), a multishell diffusion sequence not analyzed in the present study [13]. In the present study, all subjects with usable survival information in the publicly accessible release were considered, irrespective of time from surgery. This broader inclusion criterion yielded a larger usable survival subset (*n* = 133) than the 90-day RSI-anchored subset (*n* = 94) described in the original dataset publication [13], reflecting relaxation of the post-surgical timing restriction, removal of the RSI-modality eligibility criterion (not applicable to the present conventional imaging analysis), and use of the corrected public clinical follow-up release. Overall survival was defined as the interval in days from MRI acquisition to confirmed death. Subjects without a recorded date of death were right-censored at the date of last documented follow-up if a positive follow-up interval was available.

Of the 184 studies from 136 subjects in the accessible imaging release, 3 subjects lacked usable survival information and were excluded, leaving 179 scans from 133 subjects with usable survival data. For the primary analysis, only the earliest available post-treatment MRI per subject was retained in order to preserve subject-level independence and provide a single scan-anchored prognostic snapshot per patient. This yielded a first-timepoint cohort of 133 subjects comprising 110 deaths and 23 censored observations. Because the primary objective was subject-level prognostic comparison rather than longitudinal modeling, repeated follow-up scans were not included in the primary analysis.

### 2.3. Image Processing, Feature Definition, and Clinical Covariates

Images were analyzed in the preprocessed form released with the dataset, in which modalities had been co-registered, skull-stripped, and resampled to 1 mm isotropic resolution in MNI space [13]. Analysis was restricted to features derived from the neuroradiologist-approved total cellular tumor segmentation in order to maintain a single anatomically consistent tumor-mask region of interest across modalities. The total cellular tumor segmentation comprised both enhancing and non-enhancing cellular tumor components, as defined in the original dataset annotation protocol, and did not separately isolate peritumoral FLAIR hyperintensity or the surgical resection cavity [13].

Exploratory screening was performed across conventional diffusion- and perfusion-derived tumor-mask summaries available in the public release. Five summary statistics were available per map within the tumor mask (mean, median, standard deviation, 10th percentile, and 90th percentile) for *ADC*_4000_ and for the DSC-derived cerebral blood volume, cerebral blood flow, and mean transit time maps, yielding 20 conventional imaging features in total. For the focused comparison, candidate representative diffusion and perfusion heterogeneity summaries included the standard deviation of the high b-value (*b* = 4000 s/mm²) apparent diffusion coefficient within the tumor mask (*ADC*_std_) and the standard deviation of DSC-derived mean transit time within the tumor mask (*MTT*_std_); final selection of the representative features is described in Section 2.4.

Clinical covariates comprised age at MRI, sex, extent of surgical resection, and receipt of radiation therapy. MGMT promoter methylation status was analyzed separately in exploratory complete-case analyses because molecular annotation was incomplete in the public clinical release. Sex was analyzed as recorded in the source clinical dataset; gender-related variables were not available.

### 2.4. Descriptive and Exploratory Analyses

Descriptive statistics were calculated for the primary first-timepoint cohort, including event rates, censoring, and missingness of clinical and imaging variables. Because MGMT methylation status was unavailable for a substantial subset of subjects, a missingness comparison between MGMT-available and MGMT-unavailable cases was additionally performed using Fisher’s exact test for binary variables and the Mann-Whitney U test for continuous variables.

Exploratory univariate Cox proportional hazards regression was performed for the 20 conventional imaging summaries and the principal clinical covariates in order to characterize feature-level associations with overall survival. Continuous predictors were standardized prior to modeling so that hazard ratios were expressed per standard deviation increase.

Benjamini-Hochberg false discovery rate correction was applied across imaging-feature univariate *p*-values. For the focused benchmark, the diffusion and perfusion summaries exhibiting the highest univariate discrimination in the primary cohort (*ADC*_std_ and *MTT*_std_) were carried forward as the modality-level summaries with the strongest exploratory univariate signal and were subsequently used as the representative features in all incremental model comparisons. Because this feature-selection step was data-informed, benchmark results were interpreted as comparative and hypothesis-generating rather than fully confirmatory.

### 2.5. Survival Modeling and Cross-Validated Benchmark

To evaluate adjusted prognostic associations, four multivariable Cox models were fitted: clinical-only, clinical plus *ADC*_std_, clinical plus *MTT*_std_, and clinical plus both imaging features. The clinical baseline model included age at MRI, sex, extent of resection, and radiation therapy. An additional complete-case exploratory model further included MGMT methylation status. For multivariable Cox analyses, models were fitted on complete cases for the variables included in each model. Proportional hazards assumptions were assessed for the three primary adjusted Cox models (clinical plus *ADC*_std_, clinical plus *MTT*_std_, and clinical plus both imaging features) using Schoenfeld residual-based tests with rank time transformation.

To compare predictive discrimination, six models were evaluated in the cross-validation benchmark: clinical-only, diffusion-only, perfusion-only, and three clinical-incremental combinations (clinical plus *ADC*_std_, clinical plus *MTT*_std_, and clinical plus both imaging features). Five-fold stratified cross-validation was then performed in the first-timepoint cohort, with stratification based on event status [15]. Within each training fold, missing predictor values were imputed by the median and features were scaled using a robust scaler fitted on the training data only, with the same parameters then applied to the held-out validation fold. Ridge-penalized Cox models were fitted using a fixed penalizer of 0.1. Discrimination was quantified by the out-of-fold concordance index, and 1000 bootstrap resamples were used to estimate confidence intervals [16].

Predefined pairwise model contrasts were assessed using permutation testing with 2000 permutations [17]. The primary contrasts of interest were clinical plus *ADC*_std_ versus clinical-only, clinical plus *MTT*_std_ versus clinical-only, clinical plus both imaging features versus clinical-only, and diffusion-only versus perfusion-only. Additional predefined contrasts among the incremented clinical models were also examined. Because the pairwise contrasts were defined prior to the cross-validation analysis and were limited in number, multiplicity correction was not applied across these contrasts. Feature selection from the screening step was exploratory and data-informed; conclusions were therefore regarded as hypothesis-generating rather than fully confirmatory [18]. Finally, penalizer sensitivity was assessed across ridge penalties of 0.01, 0.1, and 1.0 to evaluate stability. To assess the impact of data-informed feature selection on the cross-validated benchmark, a nested sensitivity analysis was additionally performed in which the representative diffusion and perfusion features were re-selected within each training fold using the same univariate screening procedure, with discrimination evaluated on the corresponding held-out fold. To assess whether non-random MGMT missingness influenced the primary ADCstd association, a further sensitivity analysis refit the clinical-plus-*ADC*_std_ model in the MGMT-complete subset without MGMT as a covariate. To evaluate potential confounding by scan timing, a final sensitivity analysis additionally adjusted the clinical-plus-*ADC*_std_ model for log-transformed time from surgery to MRI in the subset of subjects with complete timing data.

All analyses were performed in Python 3.12 using lifelines, scikit-learn, NumPy, pandas, SciPy, and statsmodels. A two-sided p-value below 0.05 was considered statistically significant.

## 3. Results

### 3.1. Cohort Characteristics

After exclusion of scans without usable survival information, 179 scans from 133 subjects remained in the accessible release. The primary first-timepoint cohort therefore comprised 133 subjects, including 110 deaths (82.7%) and 23 censored observations (17.3%). Median overall survival among subjects with observed death events was 440 days (interquartile range, 221-650 days). Mean age at MRI was 56.5 ± 13.2 years, and 99 of 133 subjects (74.4%) were male. Gross total resection had been performed in 63 subjects (47.4%), and 58 (43.6%) had received radiation therapy. MGMT methylation status was available in 73 subjects, of whom 33 (45.2%) were methylated. Missingness affected 20 subjects (15.0%) for *ADC*_std_ and 23 subjects (17.3%) for *MTT*_std_ (Table 1).

**Table 1.** Cohort characteristics of the primary first-timepoint post-treatment glioblastoma cohort.

| Variable | Value |
| --- | --- |
| *n* | 133 |
| Deaths (events) | 110 (82.7%) |
| Censored | 23 (17.3%) |
| Median overall survival, days [IQR]* | 440 [221-650] |
| Age at MRI, years, mean ± SD | 56.5 ± 13.2 |
| Male sex, *n* (%) | 99/133 (74.4%) |
| Gross total resection, *n* (%) | 63/133 (47.4%) |
| Radiation therapy, *n* (%) | 58/133 (43.6%) |
| MGMT methylated, n/n tested (%) | 33/73 (45.2%); 60 missing |
| ADC standard deviation, mean ± SD | 0.0721 ± 0.0249 $\times 10^{-3}$ mm<sup>2</sup>/s; 20 missing |
| MTT standard deviation, mean ± SD | 1.2657 ± 0.7433 s; 23 missing |

*Note.* Median overall survival was calculated among subjects with observed death events.

Because MGMT status was unavailable in 60 subjects (45.1%), a missingness comparison was performed. Compared with MGMT-unavailable cases, MGMT-available cases were older (mean, 58.4 vs. 54.2 years; *p* = 0.043), more likely to have undergone gross total resection (57.5% vs. 35.0%; *p* = 0.014), had different *ADC*_std_ distributions (*p* = 0.012), and had longer observed survival intervals (mean, 601.1 vs. 461.1 days; *p* = 0.019). Radiation therapy also showed a borderline difference between groups (35.6% vs. 53.3%; *p* = 0.053). By contrast, no significant difference was observed for sex, event status, or *MTT*_std_. These findings indicate that the MGMT-complete subset was not missing completely at random and was therefore treated as exploratory only.

### 3.2. Exploratory Univariate Associations and Feature Selection

Exploratory univariate Cox regression across the 20 conventional imaging summaries showed that diffusion-derived heterogeneity features carried the strongest signal in the primary cohort. Among all screened imaging summaries, *ADC*_std_ demonstrated the strongest univariate prognostic association, with a hazard ratio (HR) of 1.56 per standard deviation increase, false discovery rate-adjusted *q* = 0.0003, and concordance index 0.621 (95% CI, 0.558-0.682). Additional ADC-derived summaries, including ADC_p90, ADC_median, and ADC_mean, also showed significant or near-significant associations after false discovery rate correction.

Among perfusion-derived features, *MTT*_std_ was the strongest summary, with HR 1.38, *q* = 0.0250, and concordance index 0.578 (95% CI, 0.510-0.643). MTT_p90 also remained significant after false discovery rate correction, whereas CBV- and CBF-derived summaries showed weaker and nonsignificant associations (Table 2). Among the principal clinical covariates, no variable reached conventional statistical significance in univariate Cox analysis, although gross total resection showed a trend toward lower hazard (HR 0.84, *p* = 0.067). Based on the exploratory screening step, *ADC*_std_ and *MTT*_std_ were selected as the representative diffusion and perfusion heterogeneity features for the focused multivariable and cross-validated analyses. Survival stratification based on the median split of these features showed separation between risk groups (Figure 1).

**Table 2.** Key univariate Cox proportional hazards results in the primary cohort.

| Feature | *n* | Events | HR (95% CI) | *p* | FDR *q* | C-index (95% CI) |
| --- | --- | --- | --- | --- | --- | --- |
| ADC SD | 113 | 99 | 1.56 (1.28-1.92) | < 0.001 | 0.0003 | 0.621 (0.558-0.682) |
| ADC p90 | 113 | 99 | 1.33 (1.09-1.63) | 0.0043 | 0.0290 | 0.578 (0.510-0.646) |
| ADC median | 113 | 99 | 1.31 (1.07-1.60) | 0.0081 | 0.0353 | 0.571 (0.502-0.637) |
| ADC mean | 113 | 99 | 1.27 (1.05-1.55) | 0.015 | 0.0502 | 0.563 (0.496-0.628) |
| MTT SD | 110 | 96 | 1.38 (1.12-1.69) | 0.0025 | 0.0250 | 0.578 (0.510-0.643) |
| MTT p90 | 110 | 96 | 1.33 (1.07-1.65) | 0.0088 | 0.0353 | 0.522 (0.447-0.594) |
| Age | 133 | 110 | 1.16 (0.96-1.41) | 0.1320 | — | 0.535 (0.478-0.599) |
| Male sex | 133 | 110 | 1.04 (0.86-1.27) | 0.6560 | — | 0.511 (0.465-0.558) |
| Gross total resection | 133 | 110 | 0.84 (0.69-1.01) | 0.0670 | — | 0.438 (0.386-0.491) |
| Radiation therapy | 133 | 110 | 0.88 (0.73-1.07) | 0.2030 | — | 0.477 (0.418-0.532) |

*Note.* Hazard ratios are reported per 1-standard deviation increase for continuous variables. False discovery rate correction was applied to imaging features only.

**Figure 1.**
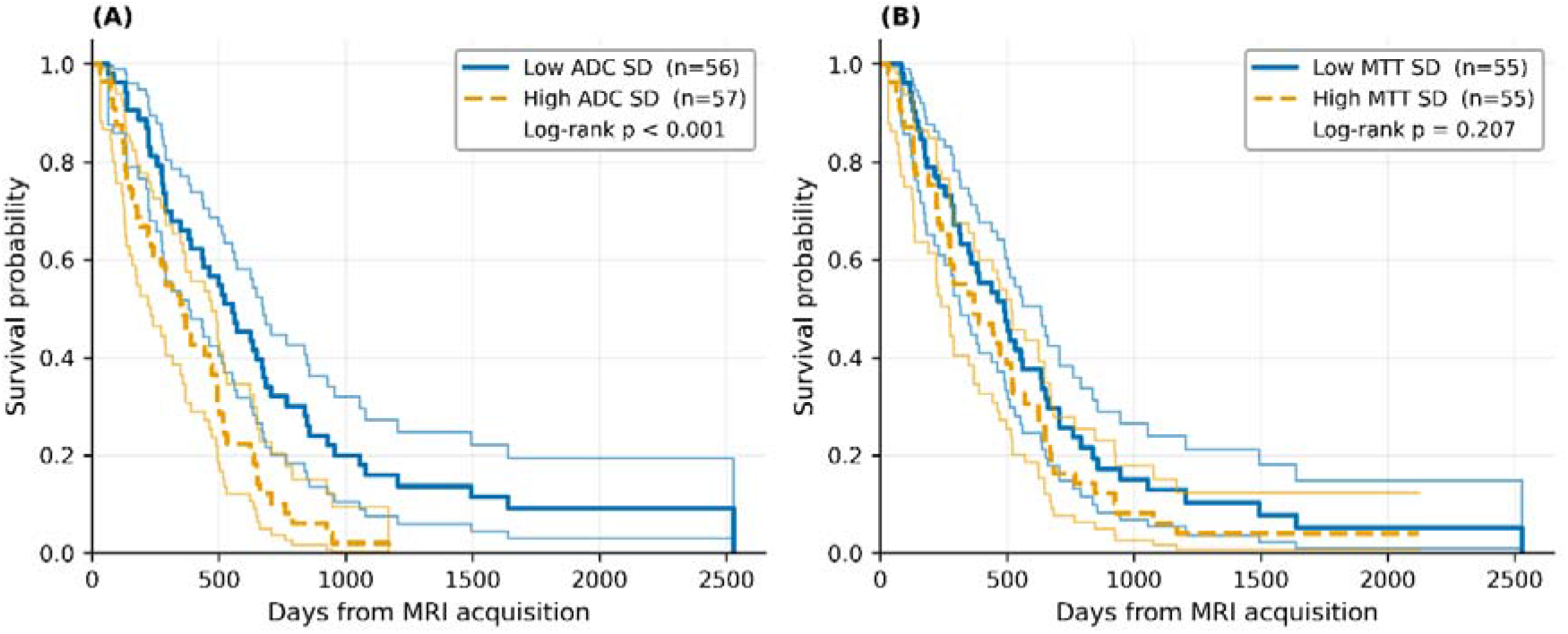
Kaplan-Meier survival curves stratified by median split of tumor-mask heterogeneity metrics in the primary first-timepoint cohort. **(A)** ADC standard deviation (ADC SD). **(B)** MTT standard deviation (MTT SD). Subjects were dichotomized into low and high groups using the cohort median for each metric. Shaded bands indicate 95% confidence intervals. Log-rank *p*-values are shown within the figure.

### 3.3. Adjusted Cox Models

In the clinical-only Cox model, none of the baseline covariates reached statistical significance, although gross total resection again showed a nonsignificant protective trend (HR 0.85, *p* = 0.084). In the model adjusted for age, sex, extent of resection, and radiation therapy, *ADC*_std_ remained independently associated with worse overall survival (HR 1.48, 95% CI 1.21-1.82, *p* < 0.001; *n* = 113, 99 events). In the corresponding clinical plus MTT model, *MTT*_std_ also remained significantly associated with worse survival (HR 1.31, 95% CI 1.07-1.60, *p* = 0.0082; *n* = 110, 96 events) (Table 3).

**Table 3.** Adjusted Cox results for the principal imaging features and corresponding model discrimination.

| Model | *n* (events) | Imaging feature | Adjusted HR (95% CI) | *p* | Apparent C | Out-of-fold C-index (95% CI) |
| --- | --- | --- | --- | --- | --- | --- |
| Clinical + ADC | 113 (99) | ADC SD | 1.48 (1.21-1.82) | < 0.001 | 0.629 | 0.605 (0.546-0.668) |
| Clinical + MTT | 110 (96) | MTT SD | 1.31 (1.07-1.60) | 0.0082 | 0.609 | 0.582 (0.516-0.646) |
| Clinical + Both | 110 (96) | ADC SD | 1.43 (1.16-1.77) | 0.001 | 0.639 | 0.619 (0.561-0.678) |
| Clinical + Both | 110 (96) | MTT SD | 1.23 (0.99-1.52) | 0.058 | 0.639 | 0.619 (0.561-0.678) |

*Note.* Adjusted Cox models included age at MRI, sex, extent of resection, and radiation therapy. Apparent C denotes in-sample concordance and should be interpreted as optimistic. Cross-validated C-indices are out-of-fold estimates with bootstrap 95% confidence intervals. In the clinical-only model (apparent C = 0.593; out-of-fold C = 0.547), gross total resection showed a nonsignificant protective trend (HR 0.85, 95% CI 0.69–1.01, *p* = 0.084); no other clinical covariate reached significance in any model (data not shown). In the joint model (“Clinical + Both”), ADC SD and MTT SD are reported in separate rows because both imaging features were entered simultaneously into the same Cox model.

When both imaging features were entered jointly with the clinical covariates, *ADC*_std_ retained statistical significance (HR 1.43, 95% CI 1.16-1.77, *p* < 0.001), whereas *MTT*_std_ was attenuated to borderline significance (HR 1.23, 95% CI 0.99-1.52, *p* = 0.058) (Figure 2). Apparent concordance indices were 0.593 for the clinical-only model, 0.629 for clinical plus ADC, 0.609 for clinical plus MTT, and 0.639 for clinical plus both imaging features.

**Figure 2.**
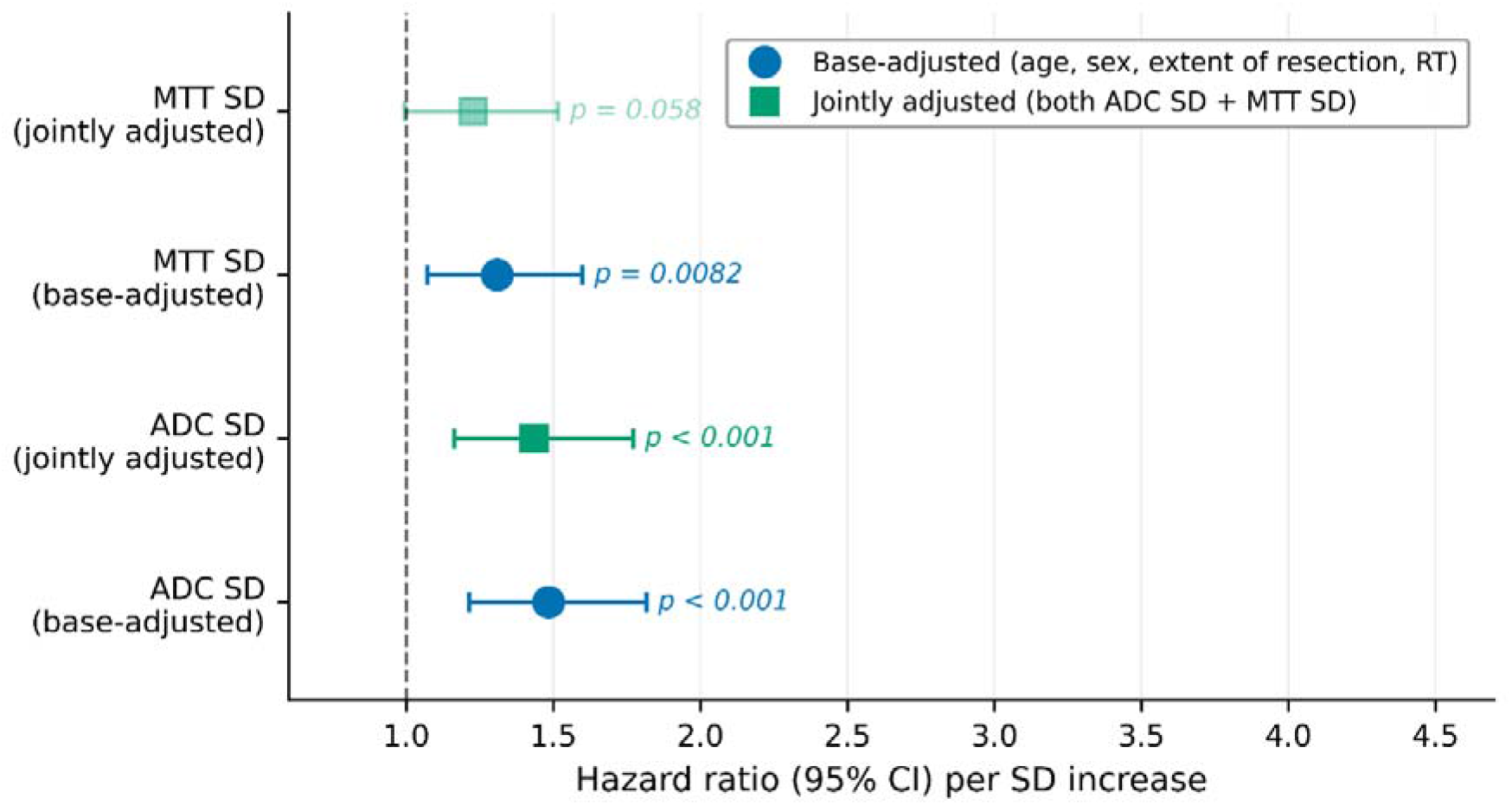
Forest plot of multivariable Cox proportional hazards models for the principal imaging features. Hazard ratios and 95% confidence intervals are shown for ADC SD and MTT SD in base-adjusted models and in the jointly adjusted model including both imaging features. Base-adjusted models included age, sex, extent of resection, and radiation therapy. Jointly adjusted models included both ADC SD and MTT SD in addition to the same clinical covariates.

*ADC*_std_ and *MTT*_std_ showed low correlation in the primary cohort (Spearman *r* = 0.24, *p* = 0.012; *n* = 110) (Figure S1), supporting their interpretation as weakly overlapping rather than strongly redundant summaries. Proportional hazards assumptions were not violated in the three primary adjusted Cox models (global Schoenfeld residual-based tests all *p* > 0.05).

Time from surgery to MRI was available for 105 of 133 subjects (median 81 days, IQR 33-302) and showed a low correlation with *ADC*_std_ (Spearman *r* = 0.30, *p* = 0.0054, *n* = 87). In a sensitivity analysis additionally adjusting for log-transformed scan timing in the complete-case subset (*n* = 87, 75 events), *ADC*_std_ remained independently associated with overall survival (HR = 1.32, 95% CI 1.04-1.67, *p* = 0.021), with timing itself showing no independent association (HR = 1.07, 95% CI 0.77-1.50, *p* = 0.674), indicating that the primary association was not substantially confounded by scan timing.

### 3.4. Cross-Validated Discrimination Benchmark

In five-fold cross-validation, the clinical-only model yielded an out-of-fold concordance index of 0.547 (95% CI, 0.480-0.614). Diffusion-only modeling based on *ADC*_std_ reached 0.605 (95% CI, 0.546-0.665), whereas perfusion-only modeling based on *MTT*_std_ reached 0.567 (95% CI, 0.503-0.627). The clinical plus ADC model also yielded a concordance index of 0.605 (95% CI, 0.546-0.668), the clinical plus MTT model 0.582 (95% CI, 0.516-0.646), and the clinical plus both imaging features model 0.619 (95% CI, 0.561-0.678), which was the highest overall discrimination observed (Figure 3; Table S1).

**Figure 3.**
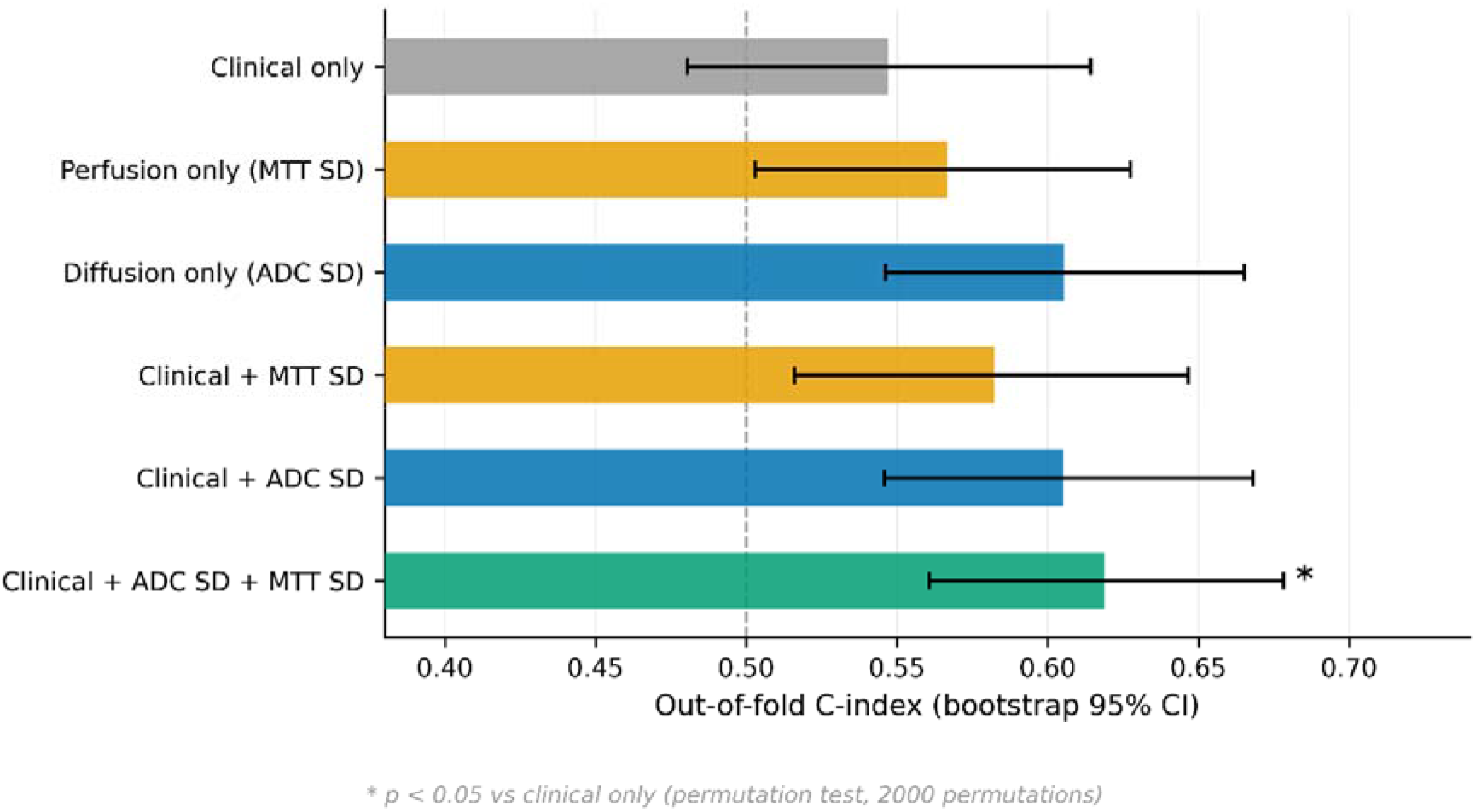
Cross-validated discrimination benchmark in the primary first-timepoint cohort. Bars represent out-of-fold concordance indices for the evaluated models: clinical-only, diffusion-only (ADC SD), perfusion-only (MTT SD), and their incremental combinations. Error bars indicate bootstrap 95% confidence intervals (1000 resamples). The asterisk (*) denotes *p* < 0.05 versus the clinical-only model by permutation testing (2000 permutations).

Permutation testing showed that the clinical plus both imaging features model significantly outperformed the clinical-only baseline (ΔC = +0.0718, *p* = 0.029). By contrast, the improvement for clinical plus ADC over clinical-only was borderline (ΔC = +0.0582, *p* = 0.071), whereas clinical plus MTT did not significantly improve discrimination over the clinical baseline (ΔC = +0.0354, *p* = 0.194). Direct diffusion-only versus perfusion-only comparison also did not reach significance (ΔC = +0.0388, *p* = 0.307), although the point estimate favored diffusion. Among additional pairwise comparisons, clinical plus both did not differ significantly from clinical plus ADC (ΔC = +0.0136, *p* = 0.314), whereas clinical plus both showed a trend toward improved discrimination over clinical plus MTT (ΔC = +0.0364, *p* = 0.086), consistent with ADC heterogeneity contributing the numerically stronger incremental signal. Penalizer sensitivity analysis showed stable model ranking across ridge penalties of 0.01, 0.1, and 1.0, supporting that the observed performance ordering was not driven by the specific penalization strength. In a nested cross-validation sensitivity analysis, *ADC*_std_ and *MTT*_std_ were re-selected as the representative diffusion and perfusion features in all five folds; nested out-of-fold concordance indices were 0.605 (95% CI, 0.538-0.667) for the diffusion-only model and 0.567 (95% CI, 0.509-0.624) for the perfusion-only model, virtually identical to the corresponding pre-selection estimates (Table S2).

### 3.5. Exploratory MGMT Complete-Case Analysis

In the complete-case subset with available MGMT status and both imaging features (*n* = 61; 55 events), methylated tumors showed longer median overall survival than unmethylated tumors (574 vs. 444 days; log-rank *p* = 0.026) (Figure S2). In univariate complete-case exploratory analyses, both *ADC*_std_ (HR 1.33, 95% CI 1.02-1.73, *p* = 0.034) and *MTT*_std_ (HR 1.37, 95% CI 1.04-1.81, *p* = 0.025) remained associated with worse survival. However, in the multivariable exploratory model including both imaging features and MGMT, none of *ADC*_std_, *MTT*_std_, or MGMT reached statistical significance (*ADC*_std_ *p* = 0.188; *MTT*_std_ *p* = 0.250; MGMT *p* = 0.061). Given the substantial and non-random MGMT missingness, these results were interpreted as exploratory only. As a further sensitivity analysis, the clinical-plus-*ADC*_std_ model was refit in the MGMT-complete subset (*n* = 62, 56 events) without MGMT as a covariate; the association of *ADC*_std_ with overall survival remained directionally consistent (HR 1.25, 95% CI 0.96-1.63, *p* = 0.096) (Table S3), with attenuation to non-significance in this smaller subset despite preservation of effect direction, consistent with reduced statistical power rather than reversal of the association.

## 4. Discussion

The principal finding of this focused benchmark was that intratumoral heterogeneity on high b-value ADC maps represented the strongest prognostic imaging signal within a parsimonious post-treatment tumor-mask framework. Among the 20 conventional imaging summaries screened, ADC standard deviation (*ADC*_std_) showed the strongest univariate association with overall survival and retained its significance after adjustment for base clinical covariates. MTT standard deviation (*MTT*_std_) emerged as the strongest perfusion-derived feature but contributed a weaker independent signal. In the cross-validated benchmark, the highest overall discrimination was achieved by combining clinical, diffusion, and perfusion features (C-index = 0.619), which was the only model to significantly outperform the clinical baseline (*p* = 0.029). Although this comparison was pre-specified, multiplicity correction was not applied across the full set of pairwise contrasts; under a conservative Bonferroni threshold for the four primary contrasts (α ≈ 0.0125), this result would not retain significance, and accordingly the advantage of the combined model over the clinical baseline should be interpreted as hypothesis-generating rather than definitive. A nested feature-selection sensitivity analysis yielded nearly identical discrimination estimates to the pre-selection benchmark outlined in Section 3.4, suggesting that residual optimism from the feature-selection step was likely limited. These findings suggest that, within this benchmark, diffusion heterogeneity contributed the numerically strongest imaging signal, while perfusion heterogeneity provided modest complementary value.

The prognostic value of ADC heterogeneity is biologically plausible in the post-treatment setting. Elevated ADC standard deviation reflects spatial complexity and microstructural disorganization within the tumor mask, driven by the coexistence of viable tumor, edema, necrosis, and treatment-related change [5]. In such a heterogeneous environment, a dispersion-based measure such as standard deviation may capture spatial variability that is not reflected by central-tendency summaries such as the mean or median. This interpretation aligns with prior studies demonstrating that imaging-derived heterogeneity adds prognostic value beyond conventional clinical models [3]. Furthermore, our findings are broadly consistent with the wider literature establishing the prognostic utility of ADC histogram-derived metrics in both newly diagnosed and recurrent, bevacizumab-treated glioblastoma cohorts, although these prior studies primarily examined the low-ADC fraction as a marker of tumor cellularity, whereas the present study identified ADC standard deviation, a heterogeneity-based summary capturing spatial dispersion rather than restricted diffusion per se, as the dominant prognostic signal [8,9]. The observed concordance indices remained in a moderate range (∼0.62), underscoring that ADC heterogeneity should be pragmatically viewed as a risk-stratifying biomarker rather than a stand-alone clinical decision tool.

The perfusion findings provide a nuanced perspective on the utility of DSC-MRI in this context. Although *MTT*_std_ was the strongest perfusion summary and remained significant in the base-adjusted Cox model, perfusion-only modeling did not significantly outperform the clinical baseline. This relatively weaker performance is technically and biologically consistent with the well-documented complexities of DSC measurements in treated GBM. In the post-treatment environment, DSC-derived signals can be substantially influenced by blood-brain barrier disruption, post-treatment vascular remodeling, and the vascular normalization effects of anti-angiogenic therapies [5,11]. Furthermore, global tumor-mask summaries may dilute localized perfusion signals and increase sensitivity to acquisition- and post-processing-related variability, including leakage correction [10,12]. The low correlation observed between *ADC*_std_ and *MTT*_std_ (Spearman *r* = 0.24), alongside the improved discrimination of the combined model, is consistent with diffusion and perfusion capturing partially distinct, non-redundant aspects of the treated tumor microenvironment.

The MGMT-related findings in this cohort require cautious interpretation. While MGMT-methylated cases exhibited significantly longer survival in the complete-case subset, consistent with established literature [2], the missingness analysis revealed systematic differences between groups. Subjects with available MGMT data were older, had higher rates of gross total resection, and exhibited systematically longer survival times than those with missing MGMT status. Consequently, the attenuation of imaging features in the MGMT-adjusted exploratory model reflects a combination of severely reduced sample size, loss of statistical power, and non-random missingness, rather than definitive evidence that the imaging effects are independent of, or fully explained by, MGMT-related tumor biology. A sensitivity analysis restricted to the MGMT-complete subset showed a directionally consistent, though no longer statistically significant, association between ADCstd and overall survival, as shown in Section 3.5, supporting that the primary ADCstd finding is unlikely to be driven solely by the missingness pattern, although reduced power in this subset precludes a stronger conclusion.

Several limitations should be acknowledged. First, all analyses were performed on a single public post-treatment cohort originating from one institutional network, which limits generalizability across differing scanners and processing environments [13]. In addition, of the 179 scans with usable survival information, 46 represented repeated follow-up examinations beyond the earliest eligible scan and were not incorporated into the present analysis, which was restricted to a single scan-anchored snapshot per subject to preserve subject-level independence. Future work could explicitly model these examinations using longitudinal or time-varying survival frameworks. Second, the representative diffusion and perfusion features were selected after exploratory screening within the same cohort. A nested sensitivity analysis re-selecting features within each cross-validation fold yielded near-identical performance estimates, as detailed in Section 3.4, suggesting limited residual optimism bias in this benchmark; nonetheless, the overall feature-selection-then-benchmarking strategy remains data-informed and the findings should be interpreted as hypothesis-generating [18]. Third, the study design was intentionally restricted to coarse, global tumor-mask summary statistics. While this approach maximizes clinical accessibility and reproducibility, it may underrepresent the prognostic potential of spatially resolved, voxel-wise, or subregion-specific imaging analyses. Additionally, the high b-value (*b* = 4000 s/mm²) ADC maps used here are not part of routine clinical diffusion protocols, which typically employ lower b-values (*b* ≈ 800-1000 s/mm²); the generalizability of the present findings to conventional clinical ADC acquisitions remains to be established. Furthermore, ADCstd as a dispersion measure is not normalized to the mean and may partly covary with mean ADC; a coefficient-of-variation-based summary was not evaluated here but represents a reasonable alternative for future work. Fourth, the scan-anchored survival definition means that subjects with a later first eligible scan have, by construction, already survived longer post-surgically at the start of follow-up; this is a structural feature of the scan-anchored design rather than a covariate that can be fully adjusted away. Relaxation of the post-surgical timing restriction used in the original dataset publication increased the usable sample size but also introduced greater heterogeneity in treatment phase, as the variable timing of MRI examinations relative to surgery and treatment may reflect different stages of disease evolution and treatment response. Time from surgery to MRI showed a low correlation with *ADC*_std_, and *ADC*_std_ remained significantly associated with overall survival after adjustment for this interval in a complete-case sensitivity analysis, suggesting that scan timing does not fully explain the primary finding. Finally, the lack of an external validation cohort with matched post-treatment diffusion and perfusion imaging precludes the direct assessment of model transportability.

## 5. Conclusion

This focused benchmark identified intratumoral heterogeneity on high b-value ADC maps as the numerically strongest prognostic imaging signal within this cohort, with DSC-derived perfusion heterogeneity contributing weaker but partially distinct information. ADC standard deviation showed the strongest univariate association among the imaging features evaluated and remained independently significant in adjusted Cox models, whereas MTT standard deviation contributed a more modest, attenuated signal. In the cross-validated benchmark, the model combining clinical variables with both ADC and MTT heterogeneity achieved the highest numerical discrimination (C-index = 0.619) and was the only model to significantly outperform the clinical baseline (ΔC = +0.072, *p* = 0.029); neither imaging feature alone reached this threshold (clinical plus ADC, *p* = 0.071; clinical plus MTT, *p* = 0.194). However, this combined model did not differ significantly from the clinical-plus-ADC model alone (*p* = 0.314), so the increment over ADC alone did not reach statistical significance, and whether MTT heterogeneity provides a genuine additional contribution beyond ADC remains an open question for future, larger cohorts. External validation in larger, independent post-treatment cohorts is warranted before these findings can inform clinical risk stratification.

## Data Availability

The imaging and clinical data analyzed in this study are publicly available through The Cancer Imaging Archive (TCIA) as part of the UCSD-PTGBM dataset (https://doi.org/10.7937/fwv2-dt74). Analysis code is available from the author upon reasonable request.

## Acknowledgements

Gratitude is extended to the researchers at the University of California San Diego (UCSD) for the generation and annotation of the UCSD-PTGBM dataset, and to The Cancer Imaging Archive (TCIA) for providing public access to these data. Furthermore, the SAIC Metadata Content Development Team is acknowledged for identifying and creating the caDSR Common Data Elements used to describe the harmonized components of this dataset. This work was supported by the CBIIT under Task Order 140D0421F0008 from the National Cancer Institute (NCI)

