## Supplemental Figures for "Diffusion and Perfusion Heterogeneity for Survival Stratification in Post-Treatment Glioblastoma"

**Supplementary Figure Legends**

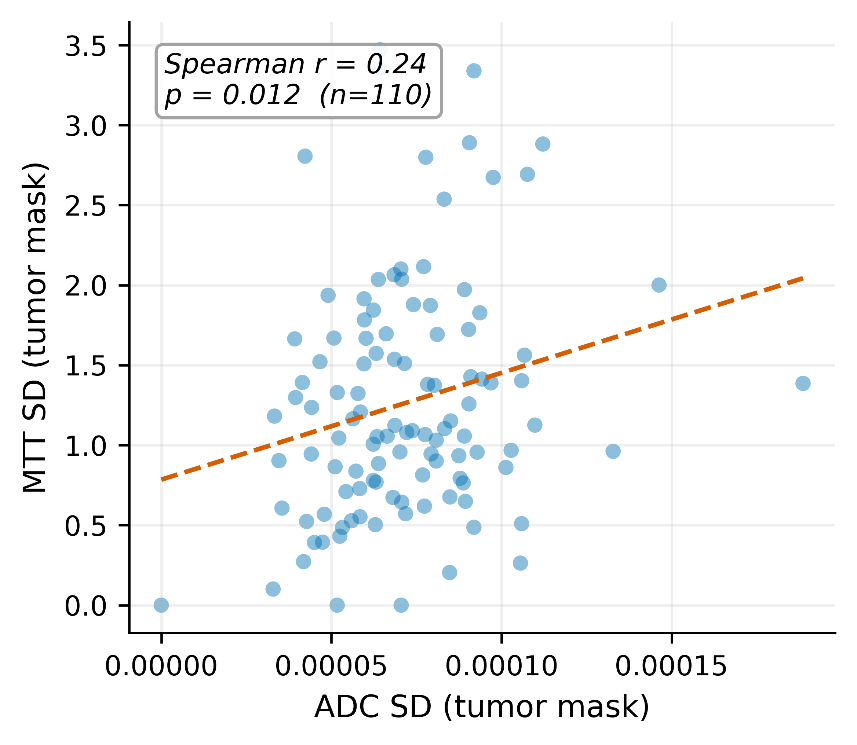

**Figure S1.** Scatter plot showing the relationship between ADC SD and MTT SD in the primary first-timepoint cohort. The dashed line represents the least-squares linear fit. Spearman correlation coefficient and *p*-value are displayed within the figure.

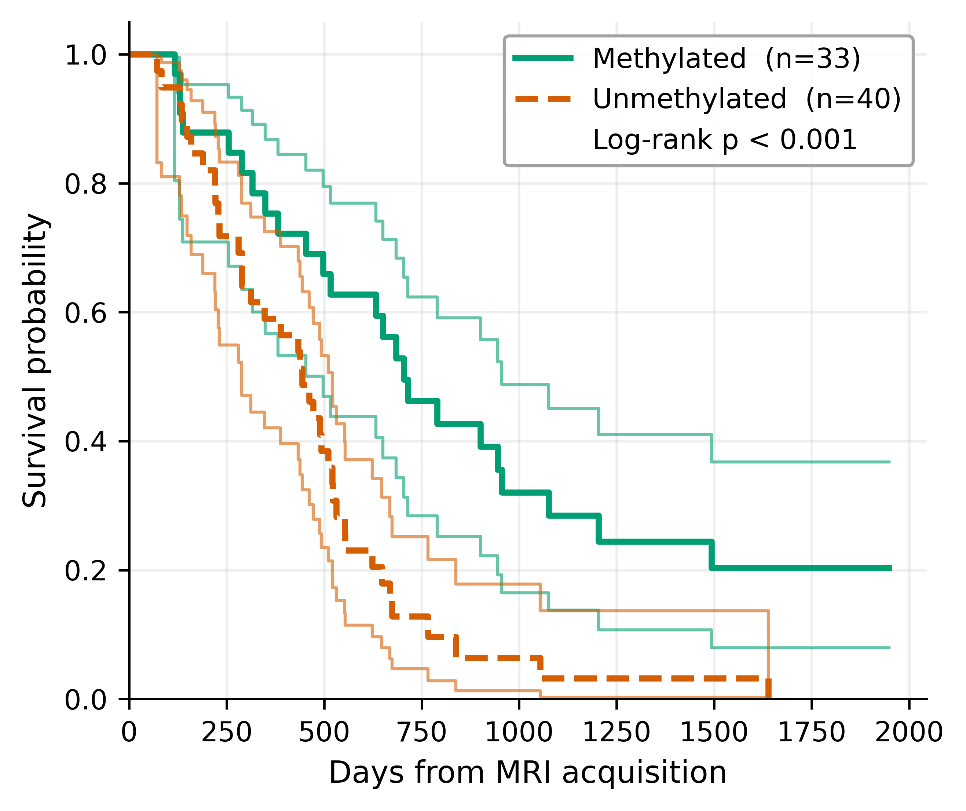

**Figure S2.** Kaplan-Meier survival curves according to MGMT methylation status in the complete-case subset with available molecular data. Shaded bands indicate confidence intervals. The log-rank *p*-value is shown within the figure. Because MGMT data were missing in a substantial and non-random subset of the cohort, this analysis was interpreted as exploratory.
