## Supplemental Tables for "Diffusion and Perfusion Heterogeneity for Survival Stratification in Post-Treatment Glioblastoma"

**Supplementary Tables**

**Table S1. Cross-validated discrimination benchmark in the primary cohort**

| **Model** | ***n* (events)** | **C-index (95% CI)** | **ΔC vs Clinical** | ***p* (permutation)** |
| --- | --- | --- | --- | --- |
| Clinical only | 133 (110) | 0.547 (0.480-0.614) | reference | — |
| Diffusion only (ADC SD) | 133 (110) | 0.605 (0.546-0.665) | reference | — |
| Perfusion only (MTT SD) | 133 (110) | 0.567 (0.503-0.627) | reference | — |
| Clinical + ADC SD | 133 (110) | 0.605 (0.546-0.668) | +0.0582 | 0.071 |
| Clinical + MTT SD | 133 (110) | 0.582 (0.516-0.646) | +0.0354 | 0.194 |
| Clinical + ADC SD + MTT SD | 133 (110) | 0.619 (0.561-0.678) | +0.0718 | 0.029* |

*Note.* C-indices are out-of-fold estimates with bootstrap 95% confidence intervals based on 1000 resamples. Permutation testing used 2000 permutations and was performed against the clinical-only model.
* p < 0.05.

**Table S2. Nested feature-selection cross-validation sensitivity analysis**

**Part A: Fold-wise feature selection**

| **Fold** | ***n* (train)** | ***n* (val)** | **Diffusion feature selected** | **Perfusion feature selected** |
| --- | --- | --- | --- | --- |
| 1 | 106 | 27 | ADC SD | MTT SD |
| 2 | 106 | 27 | ADC SD | MTT SD |
| 3 | 106 | 27 | ADC SD | MTT SD |
| 4 | 107 | 26 | ADC SD | MTT SD |
| 5 | 107 | 26 | ADC SD | MTT SD |

*Note*. ADC SD was selected in 5/5 folds and MTT SD in 5/5 folds, identical to the pre-selected representative features used in the primary benchmark.

**Part B: Nested versus pre-selected out-of-fold C-index**

| **Model** | **Pre-selected C-index (95% CI)** | **Nested C-index (95% CI)** |
| --- | --- | --- |
| Diffusion only (ADC SD) | 0.605 (0.546–0.665) | 0.605 (0.538–0.667) |
| Perfusion only (MTT SD) | 0.567 (0.503–0.627) | 0.567 (0.509–0.624) |

*Note.* Nested estimates were obtained by re-selecting the representative diffusion and perfusion features within each cross-validation training fold and evaluating discrimination on the corresponding held-out fold.

**Table S3. MGMT-complete subset sensitivity analysis**

| **Analysis** | ***n* (events)** | **HR (95% CI)** | ***p*** | **MGMT covariate** | **Note** |
| --- | --- | --- | --- | --- | --- |
| Main analysis (full cohort) | 113 (99) | 1.484 (1.212–1.816) | < 0.001 | No | All subjects with clinical and ADC SD data |
| MGMT-complete subset | 62 (56) | 1.252 (0.961–1.631) | 0.096 | No | Restricted to subjects with available MGMT data; MGMT not included as a covariate |

*Note*. ADC SD hazard ratios from the clinical-plus-ADC SD model are compared across the full cohort and the MGMT-available subset to assess whether non-random MGMT missingness confounds the primary ADC SD finding.
